# Commercially-available heart rate monitor repurposed into a 9-gram standalone device for automatic arrhythmia detection with snapshot electrocardiographic capability: a pilot validation

**DOI:** 10.1101/2021.12.16.21267762

**Authors:** Nicola Gaibazzi, Claudio Reverberi, Domenico Tuttolomondo, Bernardo Di Maria

## Abstract

**Background:** The usefulness of opportunistic arrhythmia screening strategies, using an electrocardiogram (ECG) or other methods for random “snapshot” assessments is limited by the unexpected and occasional nature of arrhythmias, leading to a high rate of missed-diagnosis.

We have previously validated a cardiac monitoring system for AF detection pairing simple consumer-grade Bluetooth low-energy (BLE) heart rate (HR) sensors with a smartphone application (RITMIA^tm^, Heart Sentinel srl, Italy). In the current study we test a significant upgrade to the abovementioned system, thanks to the technical capability of new HR sensors to run algorithms on the sensor itself and to acquire (and store on-board) single-lead ECG strips, if asked to do so.

**Methods and Result:** We have reprogrammed a HR monitor intended for sports use (Movensense HR+) to run our proprietary RITMIA algorithm code in real-time, based on RR analysis, so that if any type of arrhythmia is detected it triggers a brief retrospective recording of a single-lead ECG, providing tracings of the specific arrhythmia for later consultation.

We report the initial data on the behavior, feasibility and high diagnostic accuracy of this ultra-low weight customized device for standalone automatic arrhythmia detection and ECG recording, when several types of arrhythmias were simulated, under different baseline conditions.

**Conclusions:** The customized device was capable to detect all types of simulated arrhythmias and correctly triggered an visually interpretable ECG tracing. Future human studies are needed to address real-life accuracy of this device.

---

Technological advances in the field of cardiac rhythm monitoring have been exponential in the last years. The design of new devices or repurposed consumer technology focused mostly on the detection of silent atrial fibrillation (AF), but recent studies demonstrate limited clinical utility in the pursuit of short and asymptomatic AF episodes.[1] Still, establishing the diagnosis of several other types of brady or tachy-arrhythmias remains extremely useful, to patients and their caring cardiologists, since symptoms such as palpitations or syncope remain among the top reasons for cardiology visits. Most arrhythmias are short-lived, but the prognostic information of detecting even few seconds of some of them, for example asystole or ventricular tachycardia, is very high. The usefulness of opportunistic screening strategies, using an electrocardiogram (ECG) or other methods for random “snapshot” assessments is limited by the unexpected and occasional nature of arrhythmias, leading to a high rate of missed-diagnosis. [2]

Studies have demonstrated that prolonging the monitoring period yields incremental detection of arrhythmias [3,4], but Holter ECG recordings remain limited in the maximal duration of continuous monitoring, often not sufficient to detect rarely-occurring arrhythmias.

We have previously validated a cardiac monitoring system (for AF detection) pairing simple consumer-grade Bluetooth low-energy (BLE) heart rate (HR) sensors with a smartphone application (RITMIA^tm^, Heart Sentinel srl, Italy).[5] The application, running on a smartphone, receives beat-to-beat RR interval data by the HR sensor and applies the algorithm in real-time, generating a continuous output of either probable AF, non-AF arrhythmia or normal sinus rhythm, using a combination of RR intervals variability and chaoticity.

In the current study we aim to test a significant upgrade to the abovementioned system, thanks to the technical capability of new HR sensors to run algorithms on the sensor itself and to acquire (and store on-board) single-lead ECG strips, when asked to do so. Using an existing software development kit (SDK), we have reprogrammed a HR monitor intended for sports use (Movensense HR+) to run our proprietary RITMIA algorithm code in real-time, based on RR analysis, so that anytime any type of arrhythmia is detected it triggers a brief retrospective recording of a single-lead ECG: this provides tracings of the specific arrhythmia for later consultation.

We report the diagnostic accuracy of this affordable, ultra-low weight customized device for standalone automatic arrhythmia detection and ECG recording, when several types of arrhythmias were simulated.

## Methods

### The custom-programmed sensor

The core component of this new diagnostic system is a commercially available bluetooth low-energy (BLE) HR monitor sensor (Movesense HR+, Suunto, Finland), commercialized for use during sports, working through a chest-strap, although it can also be connected to the skin with small adhesive patch-type electrodes. The sensor is able to reliably and continuously acquire peak RR cardiac interval data [13] through the electrodes in contact with the skin. The integrated circuits filter out noise and non-cardiac electrical potentials, based on frequency or amplitude, robustly selecting and transmitting only R-R interval data in real-time, through BLE standard protocol. Importantly, this device is also capable to acquire/transmit (and store on board) single-lead electrocardiograms, if prompted to do so.

We programmed this device using the SDK optionally provided with the sensor, embedding our patented algorithm for arrhythmia detection (RITMIA^tm^, Heart Sentinel srl, Parma, Italy) so that only the robust RR interval data, in milliseconds, is used for continuous computation of variability indexes; this generates a real-time rhythm classification of “normal sinus rhythm”, “probable AF” or “undetermined non-AF arrhythmia”, this last category encompassing all events not recognized as either non-normal sinus rhythm or AF. The algorithm uses a moving data matrix comprising a number of prior R-R intervals and it measures RR heterogeneity, based on a combination of variability and chaoticity [15]. The device has been programmed to acquire 5 seconds of retrospective ECG strips if the on-board running RITMIA^tm^ algorithm recognizes an episode of undetermined non-AF arrhythmia, and 10 seconds retrospective strips at the inception and at the end of an AF episode. When the rhythm is labelled as normal sinus rhythm no ECG is recorded, since no arrhythmia is present, preserving the built-in limited memory of the device.

***The simulation environment***. Figure 1 shows the simulation environment used.

**Figure 1.**
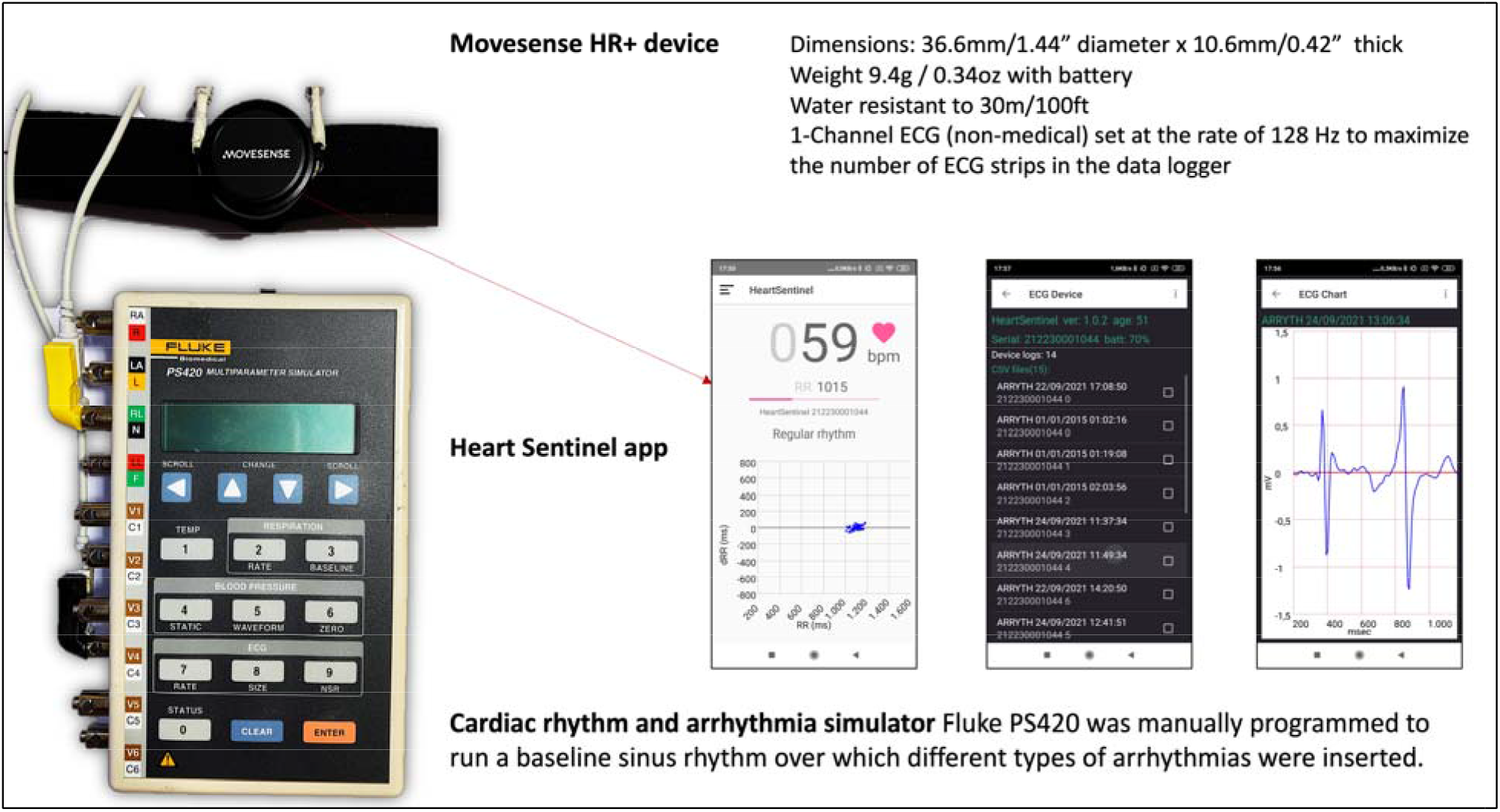
shows the simulation system components; in particular the Movesense HR+ sensor attached to the chest-belt, and in parallel wired to the output of a Fluke PS420 simulator. In the middle, three screenshots from the Heart Sentinel app are shown, from left to right demonstrating: a real-time RR plot when It is connected to the sensor, a log of the ECG strips recorded on the device and a sample of 5-second ECG strip triggered by a premature ventricular beat, obtained after clicking on one of the events in the log. The Heart Sentinel app (in this case running on an Android smartphone) was used during simulations for continuous upload of RR intervals on the cloud, to be later shown in the back-office cloud app matched with on-device stored ECG tracings.

In healthy human subjects at least some heart rate variability is present, so that for example the root mean square of the successive differences among multiple beats (RMSSD) is always higher than zero, around 0.025 in a typical healthy sitting adult; on the contrary, the use of an electronically simulated baseline sinus rhythm at a sharply fixed rate shows non-physiological absence of variability (Figure 2, Top). We conducted the arrhythmia simulation tests both using this non physiological fixed-rate sinus rhythm and also varying the rate of simulated normal sinus rhythm (Figure 2 Bottom), something that repristinates the RMSSD in the same order of magnitude of the variability of a human sitting adult.

**Figure 2.**
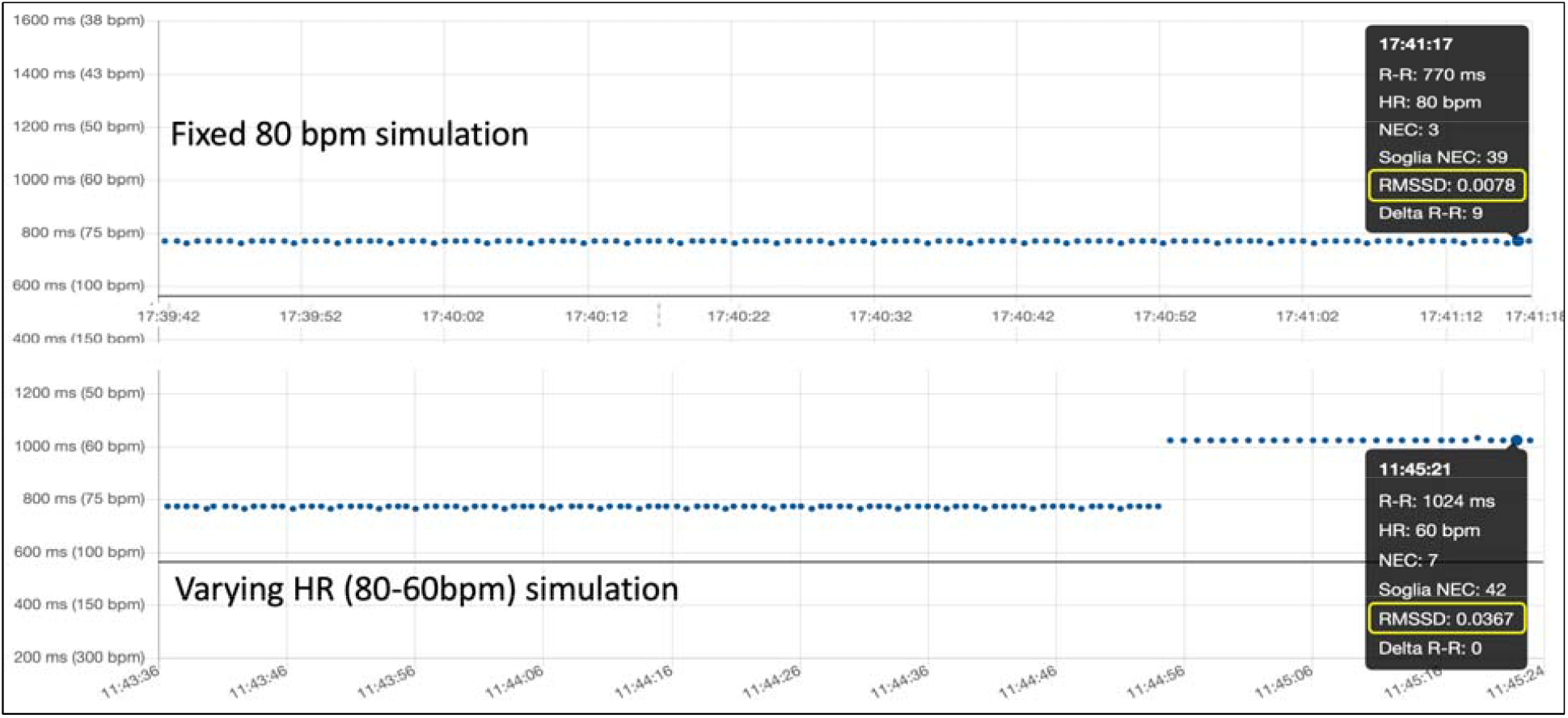
While the physiological heart rate variability usually corresponds to a RMSSD index around 0.025 in adults in the sitting position, the simulation with a fixed heart rate (Top) does not account for physiological RR variability (RMSSD 0.0078), while varying heart rate between 80bpm and 60 bpm in the simulation brings the RMSSD index back to the order of magnitude of physiological values (RMSSD 0.0367) (Bottom).

The interrogation of the device, to download the ECG strips that the device autonomously acquired (also available as comma separated value files), was conducted at the end of the arrhythmia simulations, using a BLE connected Android application (Heart sentinel application) The last ECG recording from the log was per-protocol discarded because a false alarm is always triggered by artifacts when the device is removed from the body, before it goes into standby mode. The RR intervals and their classification as either normal rhythm or one of the two abovementioned types of arrhythmias (AF or non/AF), based on the RR-based algorithm, were also available from the back-office app on the cloud, where the Heart sentinel app uploads all RR data (only if kept connected to the sensor via BLE). In this experimental setting the sensor both sends the RR intervals to the smartphone app (being uploaded to the back-office cloud app in near real-time) and at the same time it works as a fully standalone device for what concerns computing the RR algorithm and triggering the recording of ECGs (with on-board storage) if an arrhythmia is detected.

No human subjects were involved in this study.

## Results

Figure 3 shows the RR interval results of the sequential simulation of two different types of premature ventricular beats (PVCs), both of the left ventricle origin type, then of a long asystole, lasting 20 seconds, then another shorter asystole of only 7 seconds, then a single missing beat and a short (non-sustained) ventricular tachycardia. After each simulated arrhythmic event the rhythm was reverted to at least few seconds of normal sinus rhythm at 80 beats per minute. The following arrhythmia was simulated only after the rhythm was once again recognized as normal (blue labelled).

**Figure 3.**
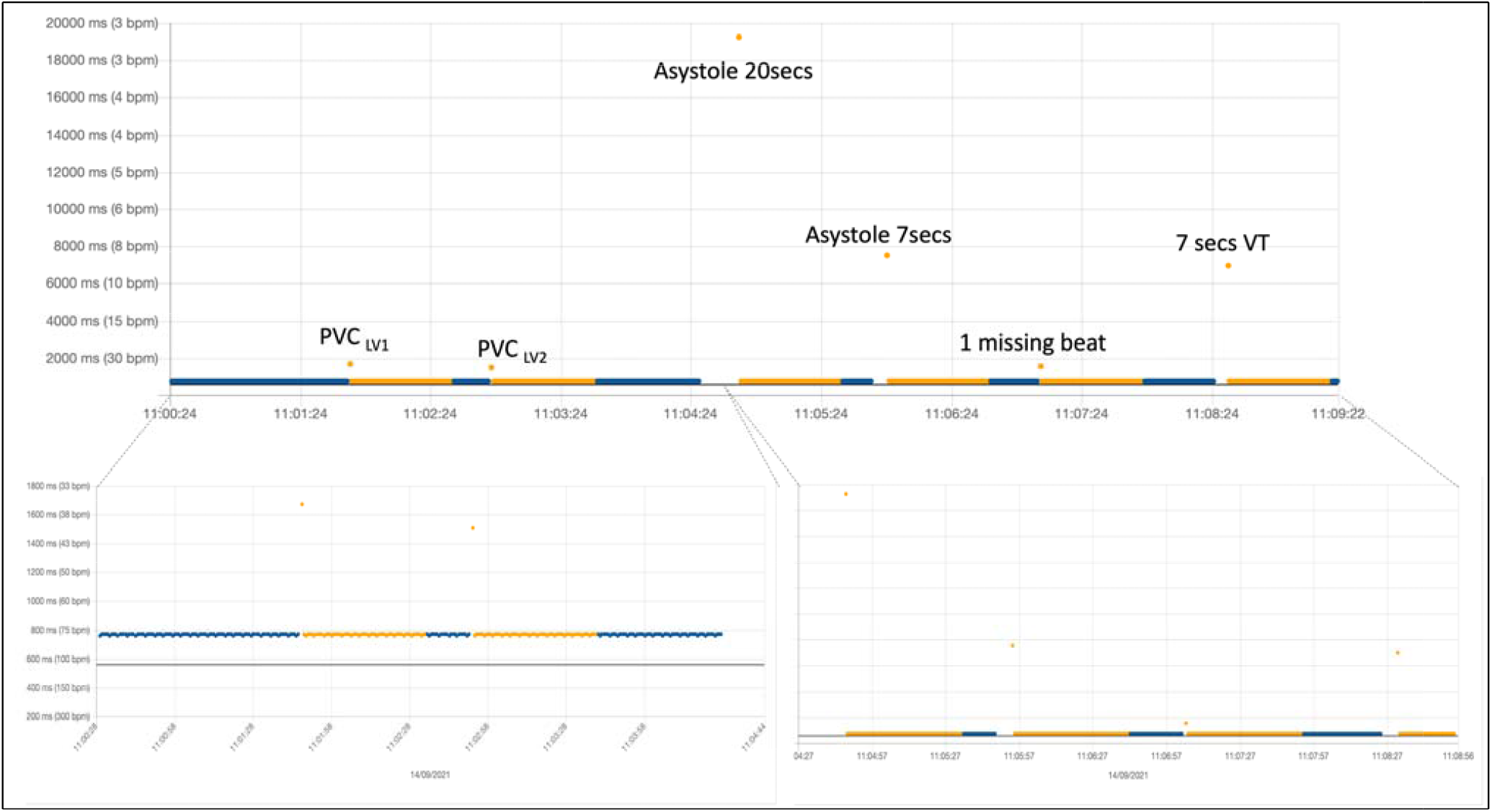
PVC, premature ventricular complex; VT, ventricular tachycardia. Beats considered from normal sinus rhythm are automatically labelled in blue, while non-AF arrhythmia in orange (and if AF is present it is labelled in red). After a single PVC or any other abnormal beat (or pause) recognized as an arrhythmic event by the algorithm, a given number of subsequent beats remain labelled in orange until the last arrhythmic beat exits the moving matrix made by a fixed number of consecutive prior beats, used for computation of variability in real-time. This orange-labelled beats always follow the arrhythmic event but they cannot trigger the acquisition of other electrocardiograms until the rhythm reverts back to normal (at least one blue beat).

Figure 4 shows the ECG tracings (5 seconds) downloaded from the device at the end of the simulation sequence shown in Figure 3. Tracings are here superimposed to the cloud-uploaded RR intervals, according to their timestamp, to match the RR abnormality that actually triggered such retrospective ECG acquisition on the device. All simulated abnormalities did effectively trigger an ECG recording, which is clearly diagnostic for the type of simulated arrhythmia. The simulation sequence was repeated 3 times and, as expected the behavior of the device did not vary.

**Figure 4.**
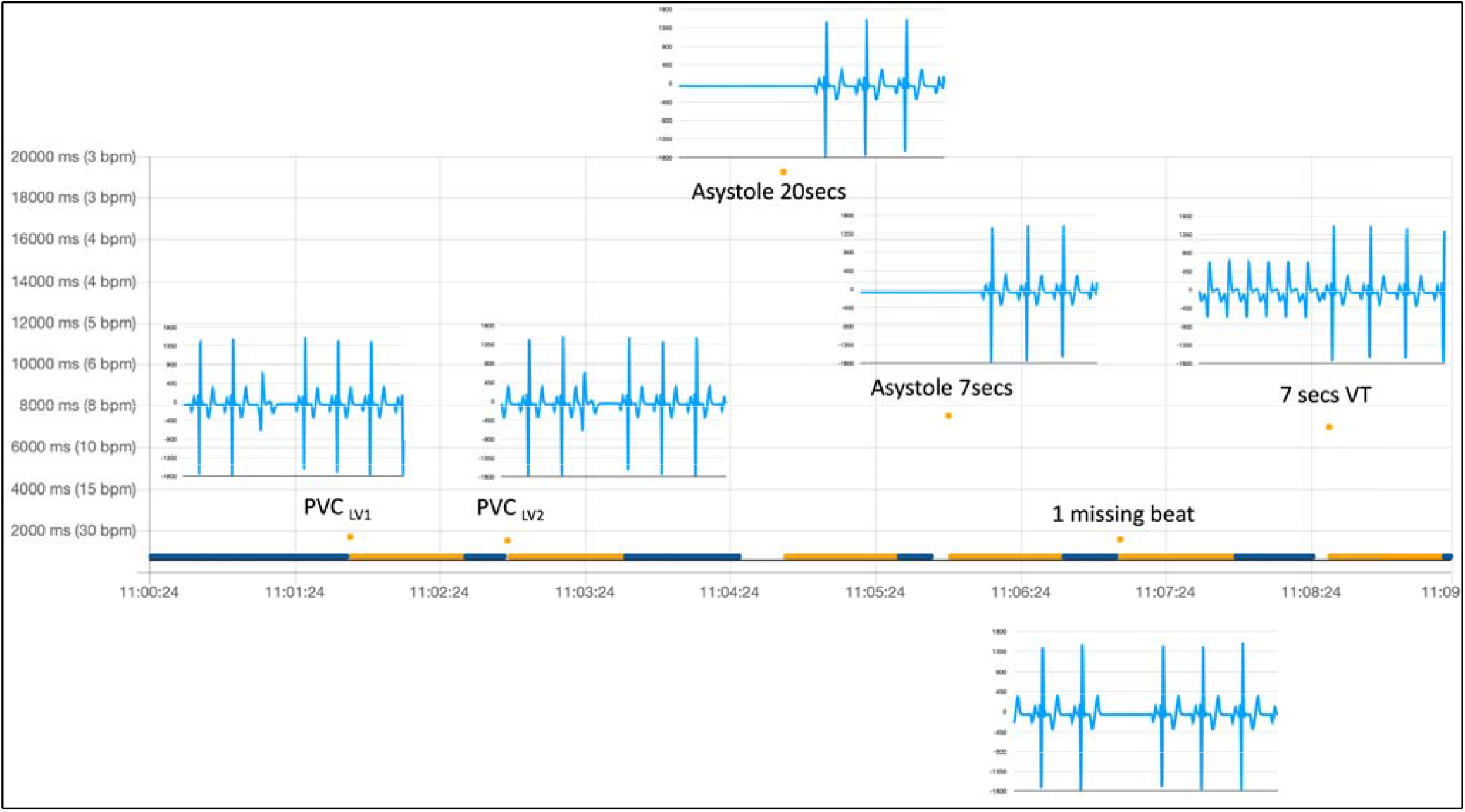
Abbreviations as in figure 3.

In figure 5 we show the same arrhythmia simulation sequence already shown in figure 3 and 4, but repeated under slightly different baseline conditions: in this case we simulated a more physiological baseline RR interval behavior during sinus rhythm preceding arrhythmia inception, by changing heart rate of the sinus rhythm from 80 bpm to 60 bpm (as explained in Figure 2) before inserting each of the arrhythmic events in the simulation. One of the parameters used in the RITMIA^tm^ diagnostic algorithm, a measure of heart rate is in fact influenced by RR variability data in the matrix of intervals preceding the event and the conduction of the simulation using the same sharp repeated RR interval, as typical of electronic simulations, is not what happens in real life, where significant beat to beat RR interval variation is present.

**Figure 5.**
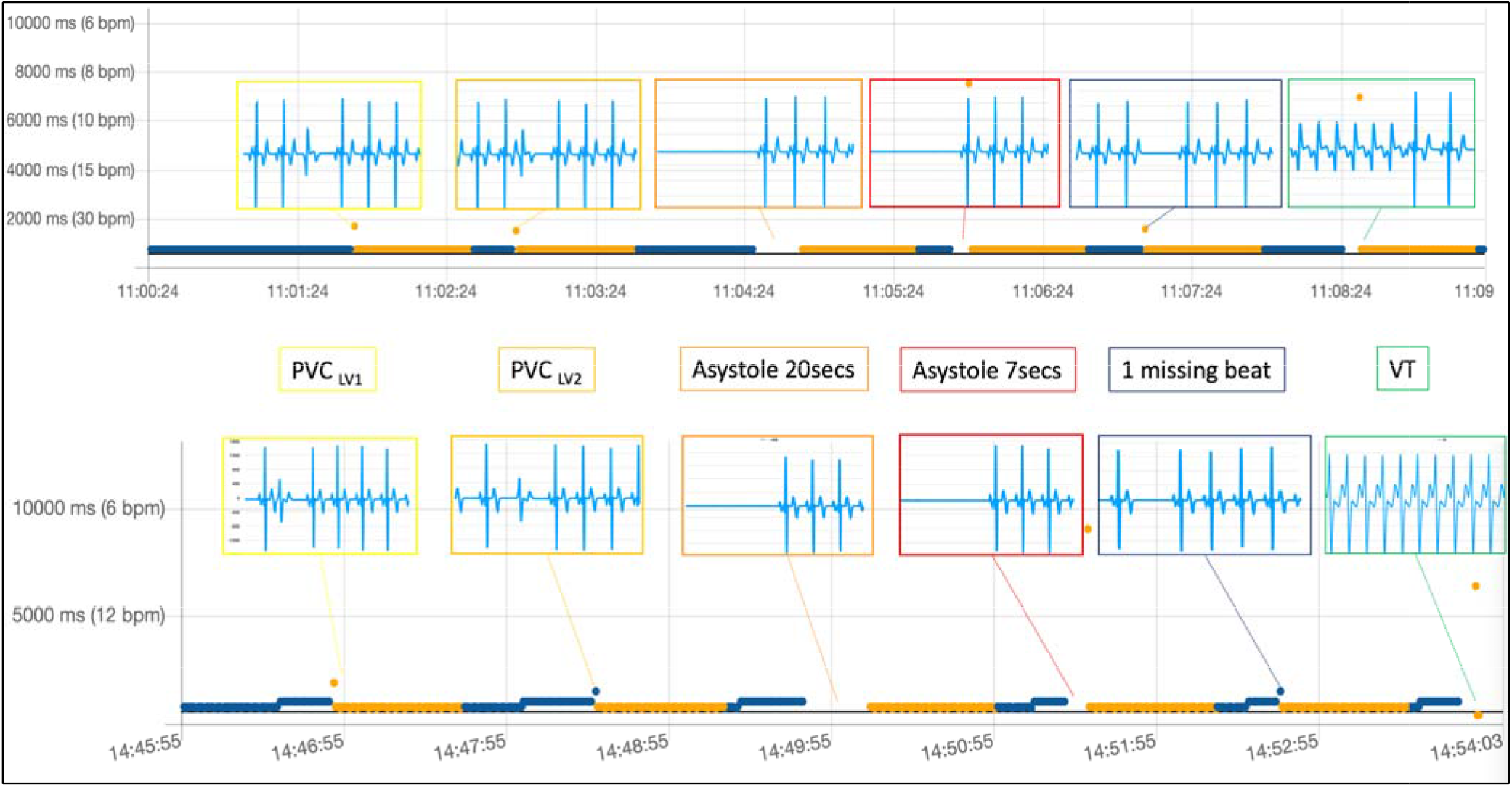
Abbreviations as in figure 3.

Figure 5 Top shows the simulation without variating the heart rate (and hence same RR intervals in the matrix preceding the arrhythmia) while at the bottom, the same sequence of arrhythmic events was simulated, but in this case only after modifying the baseline heart rate from 80 bpm to 60 bpm in the seconds preceding each simulated arrhythmia.

Results are similarly diagnostic, since all events were detected by the algorithm and the acquisition of an ECG strip was always automatically triggered, always including the main arrhythmia event in the limited 5-second duration of the ECG recordings. There are also differences, since for example “PVC LV2» and «Missing beat» were not immediately recognized as arrhythmias (see the blue color labeling of the PVC beat, meaning no arrhythmia has yet been detected) but the orange labelling comes in the immediate next beat, a different behavior compared with the fixed-rate artificial sinus rhythm at 80 bpm used in the first set of simulations reported in the top graph. This happens because using a varying heart rate as a baseline sinus rhythm, the different RR of the isolated single arrhythmic event is not sufficiently different to cause the index of RR variability cross the abnormality threshold, requiring a second interval different from the prior mean (the post-event longer RR interval) to push the variability index further towards abnormality, hence labelling this second beat as “orange” (non-AF arrhythmia) and trigger a retrospective ECG acquisition. Anytime even a single-beat arrhythmia is detected, the “orange” labelling is maintained for several seconds even if no new arrhythmia presents, because of the time needed to have the abnormal RR being excluded from the real-time matrix of many prior beats used by the algorithm to compute the RR variability.

## Discussion

New, noninvasive technologies for cardiac monitoring are flourishing; new devices, such as modified blood pressure monitors, dedicated wearable photoplethysmography-based (PPG) heart rate monitors, portable devices for single-lead ECG, traditional but smaller cabled Holter-ECG devices may all find a role in the detection of arrhythmias.

However, most of such devices have key limitations, in view of an easy applicability and patient compliance to screen long-term for arrhythmia detection: firstly, all of them require a specifically dedicated and costly hardware; secondly, most of them (with the exclusion of continuous Holter-ECG monitors) need active participation of the patient to the recording phase, reducing their utility for the detection of asymptomatic or unexpected and occasional arrhythmias; thirdly if they record in continuous, as Holter monitors do, the period they can monitor and record is limited in duration, again with less-than-optimal capability to discover occasional paroxysmal events, often taking place as rarely as few times in a year.

Rare but clinically dangerous syncopal or pre-syncopal episodes, for example, are almost impossible to be recorded with the use of external Holter monitors, although detecting asystole may signal the need for pace-maker placement; an ultralight and standalone device like the one we prototyped may be applied at the patient’s convenience anytime he prefers, even for months, with the simple caution to change the very cheap battery once in a while (once a month or less) and the comfortable adhesive patch if the chest strap is felt less comfortable.

The present feasibility and initial validation study tested a new standalone device, in its original iteration commercially available as a HR chest-belt sensor for sports. We modified its software embedding our patented arrhythmia-detection algorithms, which makes this potentially the smallest and lightest device capable to detect arrhythmias and record them in the single-lead ECG tracing format.

The cost of this native device, which is not a medical device in the HR+ model dedicated to sports, is very low, in the typical leisure-time HR monitor price range, and may remain definitely affordable for consumers once reprogrammed to become the standalone arrhythmia screening device running the RITMIA algorithm here tested.

The device is reusable, shockproof and waterproof and the coin-sized battery easily lasts more than a month, and can be easily substituted by the user (on board memory is not erased in case).

## Limitations

While non-invasive ultra-light devices are becoming increasingly useful for cardiac monitoring, they share the key limitation of limited built-in memory, so that continuous ECG recording for days or weeks is not possible, at least using the current technology and maintaining the device ultra-portable and light, which is key for patient compliance.

The way we solved this problem, without sacrificing the possibility to have ECG strips available for later cardiology consultation, is by the continuous use of our robust RR-interval algorithm to screen for the presence of a potential arrhythmia, triggering a snapshot recording of short ECG strips only in case arrhythmia is detected (thanks to the possibility to acquire retrospective ECG recordings from the data buffer). This makes the memory sufficient in this device version (a higher x300 memory version is in the pre-commercial phase) for 200-250 ECG strips, which are clinically appropriate for a device meant to screen for infrequent arrhythmias, but this mechanism fully relies on an algorithm based on RR-variability, which is reliable and robust (few false positives) but may be relatively insensitive to those rare types of arrhythmias not showing RR interval variation at their inception, during the arrhythmia or at the end.

This may be the case for example of tachycardias starting at a very similar rate compared with the preceding baseline sinus tachycardia, and not showing at least one sufficiently different RR interval at inception or the end of the arrhythmia, but this is clinically very rare, if at all possible.

## Data Availability

All data produced in the present work are contained in the manuscript.

## Notes

### Competing Interest Statement

Some of the authors (NG, CR, BDM) are part-time collaborators of the startup enterprise owning the intellectual property of the algorithms, Heartsentinel srl, Italy.
NG and CR are the inventors of the RITMIA algorithm used by the tested device.

### Funding Statement

This study did not receive any funding

